# Micropatterns of physical activity in relation to all-cause and cardiovascular disease mortality: the stealth lifestyle factor?

**DOI:** 10.1101/2024.08.06.24311529

**Authors:** Matthew N. Ahmadi, Nicholas A. Koemel, Raaj K. Biswas, Sonia Chen, Richard M. Pulsford, Mark Hamer, Emmanuel Stamatakis

## Abstract

**Importance:** Physical activity guidelines are predominantly based on questionnaire-based studies measuring only longer planned physical activity bouts (>10-15 continuous minutes). To date, short intermittent bouts of physical activity that may be beneficial for health (“micropatterns”), have counted very little towards physical activity guidelines (currently 150-300 minutes of moderate or 75-150 minutes of vigorous intensity activity per week).

**Objective:** We examined all-cause and cardiovascular disease (CVD) mortality associations of wearable device-captured activity accumulated through intermittent moderate to vigorous (IMVPA; ≤3 min) and vigorous (IVPA; ≤1 min) intensity bouts, by guideline adherence for a) active adults (eg. doing at least 150 mins/wk of moderate or 75 mins/wk of vigorous intensity physical activity), and b) inactive adults (not meeting the above amounts).

**Design:** Prospective cohort study

**Setting:** UK Biobank

**Participants:** 62,899 adults (mean age 61 years, 55.7% female) with wrist-worn accelerometer data, followed up for an average of 8.0 (SD= 0.9) years

**Exposures:** Intermittent moderate-vigorous (IMVPA; ≤3 min) and vigorous (IVPA; ≤1 min) intensity bouts; stratified by participants meeting and not meeting physical activity guidelines.

**Main outcomes and measures:** All-cause and CVD mortality obtained through linkage with the National Health Service (NHS) Digital of England and Wales or the NHS Central Register and National Records of Scotland. Cox restricted cubic spline regression were used to assess the dose-response associations.

**Results:** There were 1,583 all-cause and 442 CVD deaths among 62,899 adults (mean age 61 years, 55.7% female). Micropatterns accrued IMVPA and IVPA showed linear beneficial dose response associations in both the inactive and active groups. We observed a 1.4 to 2.0-fold lower mortality risk among inactive compared to active adults. For all-cause mortality, a median 4.0 minutes/day of IVPA was associated with a hazard ratio (HR) of 0.40 [0.32, 0.52] in inactive adults and 0.74 [0.58, 0.95] in active adults, compared to not doing any IVPA. For CVD mortality, a median of 13.0 minutes/day of IMVPA was associated with an HR of 0.32 [0.22, 0.51] in inactive adults and 0.53 [0.37, 0.78] in active adults. Analogous patterns of dose-response were present when adherence to guidelines was assessed using questionnaire-based data that only considered continuous activity bouts lasting at least 10 minutes.

**Conclusions and relevance:** Among participants not meeting guidelines, intermittent moderate-vigorous physical activity showed stronger beneficial dose-response association with all-cause and CVD mortality, compared to active adults, highlighting potential health benefits from brief activity bursts for less active adults. Considering such activity patterns are hardly represented in the evidence used to develop current guidelines, our findings support the transition of future guidelines towards wearables-informed evidence.

## INTRODUCTION

Physical activity is associated with reduced mortality and cardiovascular disease risk (CVD) ^1^. For the first time, current guidelines emphasize “all activity counts” towards accumulating the recommended 150-300 minutes of moderate physical activity or 75-150 minutes of vigorous physical activity a week. The evidence-base that shaped these guidelines comprised almost entirely of questionnaire-based studies which can only capture crude patterns of sustained physical activity bouts lasting a minimum of ≥10-15 continuous minutes ^23^. Physical activity ‘micropatterns’, defined as short bursts of intermittent physical activity lasting ≤3 minutes ^4-6^ at a time, are ignored, despite comprising >90% of adults daily physical activity^4,5^. As a consequence, micropatterns accrued activity is neither captured in the existing evidence base nor included in the weekly accumulation that contribute towards current physical activity guidelines. A 2019 review informing the most recent US guidelines concluded there was an absence of evidence suggesting activity bouts <10 minutes are not beneficial but little direct evidence for the health benefits of intermittent activity bouts was available^7^. Further, a report by the WHO Guidelines Development Group indicated the need to understand how short bursts of physical activity can influence mortality and disease risk.

Intermittent moderate to vigorous (IMVPA)^4^ and vigorous (IVPA)^5^ physical activity, can only be captured using wearable devices, such as wrist-worn trackers providing continuous high-resolution measures of movement. Small daily amounts of these traditionally “missing” activities have shown associations of sizeable magnitude with major prospective outcomes including mortality^5^, cancer incidence^6^, and major adverse cardiovascular events^4^. For questionnaire-defined inactive adults (defined as not meeting physical activity guidelines), the risk of disease and mortality is estimated to be 3-4 times greater than adults who are physically active^8,9^. Although intermittent physical activity maybe easier to accrue than prolonged exercise bouts, it is unknown if these short bouts of activity patterns confer mortality risk benefits comparable to meeting physical activity guidelines via commonly captured >10-15 minute activity bouts.

To understand the potential health benefits of intermittent physical activity among inactive and active adults, based on guideline adherence status, we examined all-cause and CVD mortality dose-response associations of wearable device-captured activity IMVPA (≤3 minute bouts) and IVPA (≤1 minute bouts).

## METHODS

### Study design and sample

Participants were included from the UK Biobank Study, a prospective cohort of 502,629 participants between 40-69 years. All participants were enrolled between 2006-2010 and provided informed written consent. Ethical approval was provided by the UK’s National Health Service, National Research Ethics Service (Ref 11/NW/0382). Participants completed physical examinations by trained staff and touchscreen questionnaires. We excluded participants with prevalent CVD, missing data on any of the assessed covariates, missing self-reported physical activity, or an event in the first 12 months of follow up (**Supplemental Figure 1**).

### Physical Activity Assessment

From 2013-2015, 103,684 participants were mailed and wore an Axivity AX3 accelerometer (Newcastle upon Tyne, UK) on their dominant wrist for 24-hrs/day for 7 days to measure physical activity. Prior to being mailed, the AX3 accelerometers were initialized to collect data with a sampling frequency of 100Hz and a dynamic range between ±8g. Participants returned the devices by mail and the data were calibrated and non-wear periods were identified according to standard procedures^10 11^. Monitoring days were considered valid if wear time was greater than 16 hours. In this study, participants were required to have at least three valid monitoring days, with at least one of those days being a weekend day. Physical activity was classified with a validated accelerometer-based machine learning physical activity classification schema, consistent with previously published studies^5 12^. We classified time spent sedentary, and in light intensity (from ambulatory activities), moderate intensity, and vigorous intensity. We further calculated time spent in IMVPA from bouts ≤3 minutes and IVPA from bouts ≤1 minute based on previous evidence regarding the health enhancing effects of intermittent activity ^4 5 13^.

Participants also self-reported physical activity using the short-form International Physical Activity Questionnaire (IPAQ) which assessed the duration and frequency of moderate, vigorous, and walking (defined as moderate activity by the IPAQ) physical activity accumulated in bouts of ≥10 continuous minutes^14^. As in previous questionnaire and wearable-based analyses^15 16^, we classified participants as physically active or physically inactive based on adherence to the lower threshold of current UK, US and WHO guidelines of 150 minutes of moderate or 75 minutes of vigorous activity a week or a combination of both.

### All-cause and cardiovascular disease mortality ascertainment

Participants were followed up through November 30th, 2022, with deaths obtained through linkage with the National Health Service (NHS) Digital of England and Wales or the NHS Central Register and National Records of Scotland. CVD was defined as diseases of the circulatory system (ICD-10 codes: I0, I11, I13, I20-I51, I60-I69), excluding hypertension, diseases of arteries, and lymph.

### Statistical Analysis

We examined the time-to-event dose-response associations between IMVPA and IVPA with all-cause and CVD mortality among active and inactive participants (defined using questionnaire and wearables-based assessments) using cox restricted cubic spline models with knots evenly spaced along the exposure distribution (eg: 10, 33, and 67 percentiles for right skewed distribution). For CVD mortality, we used Fine-gray subdistribution hazards treating non-CVD deaths as competing events. We tested the proportional hazard assumption with Schoenfeld residuals and found no evidence of violation. For each outcome, results were reported as hazard ratios (HRs) and 95% confidence intervals (CIs) with participants accumulating no IMVPA as the referent. To assess robustness of our results, we assessed the influence of the reference point by using the 10^th^ percentile of the IVPA and IMVPA daily duration distribution and provide adjusted absolute risk dose-response analyses (in person-years using Poisson regression) that is not dependent on a reference point. We further calculated the adjusted 5-year absolute risk. We report HR’s for the median daily duration IVPA of 4 mins/day (derived from full sample) and IMVPA of 13 mins/day to make direct comparisons in mortality risks among participant meeting and not meeting guidelines. To assess if IVPA or IMVPA were associated with mortality over and above meeting traditional guidelines, we performed a categorical joint association between guideline adherence and IVPA/IMVPA above and below the median duration.

We adjusted our analyses with the following covariates: age, sex, ethnicity, education, smoking status, alcohol use, fruit and vegetable consumption, family history of CVD and cancer, use of CVD medication, discretionary screen time (hours of TV and computer use), light intensity, moderate intensity (for vigorous intensity analysis), and sleep duration. In all analyses, we also included adjustment for mutually exclusive non-exposure intensity duration (e.g. in our analysis of IMVPA, we adjusted for moderate to vigorous intensity accrued from bouts lasting >3 minutes). Complete covariate definitions are provided in **Supplemental Table 1**.

In sensitivity analyses, to mitigate the potential influence of reverse causation we: 1) excluded participants with self-rated fair or poor health; 2) a body mass index <18.5 kg/m^2^; 3) high frailty index score (≥3 on a 0 to 5 scale)^17^; 4) or an event within the first 5 years of follow-up^18^. We also used a negative control outcome of deaths or hospital admission from accidents (excluding cycling, self-harm, and falls) that do not have a mechanistic link to physical activity. Negative controls can improve causal inference by illustrating pervasive bias and confounding^19^. If the negative control has a similar association pattern as the primary outcomes, then it is more plausible associations are due to bias and confounding than causal mechanisms. Because of the known associations between age and disease risk we also stratified our analyses by older adults ≥60 years based on the World Health Organization’s Healthy Ageing report^20^. We included analyses adjusting for area level socioeconomic status (Townsend Deprivation Index), in addition to individual level socioeconomic status (education – adjusted in main analysis). In additional analyses, we also adjusted for clinical factors that might be mediators (baseline waist circumference, glycated haemoglobin, HDL, LDL, and triglycerides). We used R 4.2.1 (R Core Team, 2017) and report the study per the Strengthening the Reporting of Observational Studies in Epidemiology (STROBE) guidelines.

## RESULTS

Our analytic sample included 62,968 participants (follow up= 8.0 ± 0.9 years) with an average age of 61.0 ± 7.9 years, of whom 27,876 (44.3%) were male. There was a total of 1,947 all-cause and 544 CVD deaths. Participants who met the guidelines based on wearables assessment were more likely to have never smoked (59.1% vs 55.6% never smoked status). However, the self-report assessment showed similarity in smoking history (58.2% vs 57.7% never smoking status) between those who met the guidelines and those who did not. Participants who met guidelines by either wearables or self-report assessment, on average, had a lower waist circumference than those who did not meet guidelines. **Table 1** shows the baseline sample characteristics stratified by guideline adherence for both self-report and device-based estimates. **Supplemental Figures 2-5** provide the adjusted absolute risk dose-response and 5-year mortality results. When participants accumulated less than 5 minutes/day of IMVPA, there was an average all-cause mortality risk of 20 [15, 25] deaths per 1,000 person-years for those who meeting or not meeting guidelines. When participants accumulated between 10-15 minutes/day of IMVPA, average mortality risk was 12 [8,16] for those not meeting guidelines and 15 [12, 18] for those meeting guidelines. For CVD mortality, when no IVPA was accumulated, there was an average risk of 12.5 [7.5, 20] deaths per 1,000 person-years for those meeting or not meeting guidelines. When IVPA was between 3-6 minutes/day average mortality risk was 4.4 [2.4, 5.9] deaths for those not meeting guidelines and 7 [4.8, 10.3] deaths for those meeting guidelines.

### All-cause mortality

Our all-cause mortality dose-response results showed a protective association for IMVPA and IVPA with a stronger magnitude of association for inactive participants compared to active participants (**Figure 1A-1B**). At the median 4 minutes/day of IVPA we observed an HR of 0.40 [0.32, 0.52] for inactive participants and an HR of 0.74 [0.58, 0.95] for active participants, compared to those not accruing any IVPA. For IMVPA, at the median 13 minutes/day we observed an HR of 0.42 [0.33, 0.53] for inactive participants and an HR of 0.62 [0.46, 0.84] for active participants, compared to those not accruing any IMVPA. For both groups, there was continued lower all-cause mortality risk for higher daily duration of IMVPA.

**Figure 1:** Dose-response association of all-cause mortality with physical activity accrued through micropatterns: intermittent moderate to vigorous (≤3 minute bouts), and vigorous physical activity (≤1 minute bouts) Adjusted for age, sex, ethnicity, education, smoking status, alcohol use, fruit and vegetable consumption, family history of CVD and cancer, use of CVD medication, screen time, light intensity, moderate intensity (for vigorous intensity analysis), and sleep duration, and non-exposure intensity duration (for example, vigorous activity analysis included adjustment for vigorous activity from >1 minute bouts).

### Cardiovascular disease mortality

We observed a linear beneficial dose-response association for IVPA for both active and inactive participants (**Figure 2A-B**). At the median of 4 minutes/day, CVD mortality risk among inactive participants was 0.36 [0.22, 0.60], and for active participants the risk was 0.63 [0.42, 0.94]. We also observed a stronger dose-response association for IMVPA among inactive participants compared to active participants. For inactive participants, at the median 13 minutes/day, CVD mortality risk was 0.32 [0.22, 0.51] and 0.53 [0.37, 0.78] in active adults.

**Figure 2:** Dose-response association of cardiovascular disease mortality with physical activity accrued through micropatterns: intermittent moderate to vigorous (≤3 minute bouts), and vigorous physical activity (≤1 minute bouts) Adjusted for age, sex, ethnicity, education, smoking status, alcohol use, fruit and vegetable consumption, family history of CVD and cancer, use of CVD medication, screen time, light intensity, moderate intensity (for vigorous intensity analysis), and sleep duration, and non-exposure intensity duration (for example, vigorous activity analysis included adjustment for vigorous activity from >1 minute bouts).

### Joint dose-response association of guidelines adherence

The joint IVPA-guideline adherence status analysis showed inactive adults had comparable all-cause mortality risk as active adults when IVPA daily durations reached at least 3-5 minutes (HR difference <0.01) (**Figure 3A**). For CVD mortality, the dose-response curves became nearly identical when daily durations exceeded 5 minutes/day (**Figure 4A**). The joint IMVPA-guideline adherence status analysis showed inactive adults had a comparable all-cause and CVD mortality risk as active adults when daily durations reached at least 10-15 minutes/day (HR difference <0.01) (**Figure 3B and 4B**).

**Figure 3:** Joint dose-response association of intermittent physical activity and guideline adherence with all-cause mortality. Adjusted for age, sex, ethnicity, education, smoking status, alcohol use, fruit and vegetable consumption, family history of CVD and cancer, use of CVD medication, screen time, light intensity, moderate intensity, sleep duration, and non-exposure intensity duration (for example, vigorous activity analysis included adjustment for vigorous activity from >1 minute bouts.

**Figure 4:** Joint dose-response association of intermittent physical activity and guideline adherence with cardiovascular disease mortality. Adjusted for age, sex, ethnicity, education, smoking status, alcohol use, fruit and vegetable consumption, family history of CVD and cancer, use of CVD medication, screen time, light intensity, moderate intensity, sleep duration, and non-exposure intensity duration (for example, vigorous activity analysis included adjustment for vigorous activity from >1 minute bouts.

### Self-reported classification of guidelines adherence

In the additional set of analyses where we used self-reported physical activity to determine guideline adherence, we found broadly comparable results to the main analyses above that used wearables data to categorize participants as active or inactive (**Figures 1C-D and 2C-D**). For example, 4 minutes/day of wearables-assessed IVPA was associated with an all-cause mortality risk of 0.43 [0.27, 0.59] for inactive adults and 0.55 [0.45, 0.68] for active adults. For CVD mortality, the associated risk was 0.40 [0.28, 0.69] for inactive adults and 0.48 [0.33, 0.69] for active adults.

### Sensitivity analyses

Sensitivity analyses excluding participants with self-rated fair or poor health, low body mass index, high frailty, or an event in the first 5 years of follow-up showed generally consistent dose-response associations as our main analysis with wider 95% CI’s due to reduced sample size (**Supplemental Figures 6-11**). Analyses of negative control outcomes indicated residual and unmeasured confounding had minimal impact on the findings (**Supplemental Figure 12**). Specifically, the HR point estimate pattern was inconsistent to the primary analyses for meeting guidelines and wide 95% CIs crossing the null line for those not meeting guidelines. Stratification by age showed consistent dose-response association patterns for vigorous intensity. For moderate to vigorous intensity a protective association was observed among older adults (≥60 years) and adults <60 years who did not meeting guidelines (**Supplemental Figures 13-14**).

Changing the reference point to the 10^th^ percentile of the distribution (∼1 minute of IVPA and ∼3.5 minutes of IMVPA) showed similar all-cause and CVD mortality risk between both groups when participants in the inactive group accumulated at least 2 minutes/day of IVPA or 4 minutes/day of IMVPA (**Supplemental Figures 15-16**). Additional adjustment for glycated hemoglobin, high and low density lipoproteins, triglycerides, and waist circumference did not materially change the observed associations (**Supplemental Figures 17-18**). Adjustment for Townsend Deprivation Index showed consistent dose-response association patterns as our main analysis **(Supplemental Figures 19 and 20**)

## Discussion

Our study, which examined the dose-response of micropatterns-accrued physical activity quantified by wearable trackers, demonstrated a significant and consistent linear dose-response association of IMVPA and IVPA with all-cause and CVD mortality among inactive adults, defined as participants who did not meet current physical activity recommendations. We observed an approximate 1.4 to 2.0 fold lower mortality risk for an equivalent amount of intermittent physical activity per day for inactive adults compared to active adults. Our findings highlight the substantively lower mortality risk that could be achieved through physical activity micropatterns, that are almost entirely missed by the predominately self-reported physical activity that has informed current guidelines and clinical practice.

Among adults meeting guidelines, 4 minutes/day of intermittent vigorous intensity was associated with a 25-35% lower mortality risk. At the same amount of vigorous intensity time, adults not meeting guidelines had a 50-65% lower mortality risk. For intermittent moderate to vigorous physical activity, higher daily accumulation was associated with lower risk in a linear fashion with a stronger magnitude of association for adults not meeting guidelines. The joint analyses further support these findings by showing that high levels of intermittently accrued vigorous or moderate-vigorous physical activity had comparable magnitude of associations in inactive and active participants when daily durations exceeded 3-5 minutes of vigorous intensity or 10-15 minutes of moderate-vigorous intensity activity. The single referent used in the joint dose-response analyses allows for a direct comparison between guideline adherence groups to identify when all-cause and CVD mortality risk for those not meeting guidelines may become similar to those who do meet guidelines. This minimizes the possibility that the differential dose-response associations in the two groups are due to differences in health status of two different referent groups (eg: stratification), adding information that allows for a common perspective of mortality risk across all groups.

The potential health benefits of micropatterns accrued physical activity to improve longevity and mitigate mortality risk, particularly for inactive adults, has been largely unrecognized in public health promotion strategies^21^. Our findings inform novel and potentially feasible alternatives to traditional strategies reliant on promoting prolonged and sustained exercise bouts, particularly for adults constrained by time or limited motivation or for whom longer duration or structured activity is otherwise challenging. Physical activity that can be accumulated intermittently or sporadically throughout day-to-day activities (e.g. stair climbing or short bursts of brisk to fast walking or intense gardening), could expand options for patient treatment and augment the effectiveness of physical activity prescriptions and lifestyle modification interventions.

For physically inactive adults, short intermittent bursts of moderate-vigorous and vigorous physical activity have been shown to improve lipid profile, cardiorespiratory fitness, vascular efficiency (flow mediated dilation; pulse wave velocity), and glucose metabolism^22 23^. It is plausible that these physiological and cardiometabolic adaptations contribute towards cardioprotective effects that led to the lower CVD mortality we observed. The potential cardioprotective benefits of short activity bursts maybe less potent among adults who are already sufficiently active, which would explain the attenuated, and in some instances non-significant, dose-response associations we observed for CVD mortality in participants who met the physical activity guidelines.

Using self-reported physical activity to assess guideline adherence showed a broadly similar patterns of results to the main analyses, despite the inherent differences between self-reported and wearable based measures (e.g. the self-reported measures are designed to only capture planned physical activity lasting for at least 10 continuous minutes) ^24^. Our findings have relevance for public health and clinical guidelines because people reporting little or no sustained physical activity bouts may be unaware that they are accruing short intermittent bouts of health-enhancing activity. With the proliferation of wearables among the population, future guidelines shifting towards wearables-informed evidence could place emphasis on making people aware that they can attain health-benefits from short activity bursts, even if they do not accrue prolonged activity bouts or consider themselves to be physically active.

### Strengths and limitations

Strengths of our study include the combination of wearable trackers collecting highly granular data and a validated machine learning based schema to detect and differentiate IMVPA and IVPA patterns. The large sample size and long follow up allowed us to reduce the risk of reverse causality by excluding participants with prevalence of major disease, self-rated fair/poor health, or high frailty, and those who had an event in the first two and five years of follow up. There was a median lag of 5.5 years between the UK Biobank baseline when covariate measurements and self-reported physical activity were taken and the accelerometry study. However, covariates and self-reported physical activity were generally stable over time^25^. For example, 80-85% of participants maintained their active/inactive status over 8 years of re-examination. Although some physical activity may not be perfectly captured by wearable trackers, such as carrying heavy shopping bags, such measurement error is likely random leading to underestimation of the ‘true’ associations with mortality. The UK Biobank had a very low response rate, although, evidence suggests that this and the subsequent unrepresentativeness to the target population does not affect estimates of physical activity with mortality^26^.

## Conclusion

We found that intermittently accrued vigorous and moderate-vigorous physical activity bouts were associated with lower all-cause and CVD mortality risk in both inactive and active adults, with the protective association more pronounced in the former group. There was continuing gains through higher daily durations in a generally consistent linear fashion. Individuals may find accruing intermittent physical activity more feasible in day-to-day activities, and less physically and time demanding than prolong and sustained physical activity bouts. Future trials should further investigate the potential of intermittently accrued physical activity as a potentially efficient alternative for inactive adults to attain physical activity health benefits. Considering such activity patterns are largely absent in the evidence used to develop current guidelines, our study provides an example for using wearable devices in combination with machine learning techniques to reveal novel physical activity patterns to inform public health and clinical practice. Our study also supports the transition of physical activity guidelines to incorporate towards wearables-informed evidence.

## Data Availability

All data produced in the present study are available upon reasonable request to the authors

## Acknowledgements

This study is funded by an Australian National Health and Medical Research Council Investigator Grant (APP 1194510). MH supported by the NIHR University College London Hospitals Biomedical Research Centre. This research was conducted using the UK Biobank Resource (application number 25813). The authors would like to thank all the participants and professionals contributing to the UK Biobank.

## REFERENCES

1. Bull FC, Al-Ansari SS, Biddle S, et al. World Health Organization 2020 guidelines on physical activity and sedentary behaviour. British Journal of Sports Medicine 2020;54(24):1451–62. doi: 10.1136/bjsports-2020-102955

2. Richard PT, Emmanuel S, Fiona CB. How can global physical activity surveillance adapt to evolving physical activity guidelines? Needs, challenges and future directions. British Journal of Sports Medicine 2020;54(24):1468. doi: 10.1136/bjsports-2020-102621

3. Hallal PC, Andersen LB, Bull FC, et al. Global physical activity level s: surveillance progress, pitfalls, and prospects. The Lancet 2012;380(9838):247–57. doi: 10.1016/S0140-6736(12)60646-1

4. Ahmadi MN, Hamer M, Gill JMR, et al. Brief bouts of device-measured intermittent lifestyle physical activity and its association with major adverse cardiovascular events and mortality in people who do not exercise: a prospective cohort study. Lancet Public Health 2023;8(10):e800–e10. doi: 10.1016/S2468-2667(23)00183-4

5. Stamatakis E, Ahmadi MN, Gill JMR, et al. Association of wearable device-measured vigorous intermittent lifestyle physical activity with mortality. Nature Medicine 2022;28:2521–29. doi: 10.1038/s41591-022-02100-x

6. Stamatakis E, Ahmadi MN, Friedenreich CM, et al. Vigorous Intermittent Lifestyle Physical Activity and Cancer Incidence Among Nonexercising Adults: The UK Biobank Accelerometry Study. JAMA Oncol 2023 doi: 10.1001/jamaoncol.2023.1830 [published Online First: 20230727]

7. Jakicic JM, Kraus WE, Powell KE, et al. Association between bout duration of physical activity and health: systematic review. Medicine and science in sports and exercise 2019;51(6):1213.

8. López-Bueno R, Ahmadi M, Stamatakis E, et al. Prospective associations of different combinations of aerobic and muscle-strengthening activity with all-cause, cardiovascular, and cancer mortality. JAMA internal medicine 2023;183(9):982–90.

9. Blond K, Brinkløv CF, Ried-Larsen M, et al. Association of high amounts of physical activity with mortality risk: a systematic review and meta-analysis. British journal of sports medicine 2020;54(20):1195–201.

10. Ahmadi MN, Nathan N, Sutherland R, et al. Non-wear or sleep? Evaluation of five non-wear detection algorithms for raw accelerometer data. Journal of Sports Sciences 2020;38(4):399–404. doi: 10.1080/02640414.2019.1703301

11. Sipos M, Paces P, Rohac J, et al. Analyses of Triaxial Accelerometer Calibration Algorithms. IEEE Sensors Journal 2012;12(5):1157–65. doi: 10.1109/jsen.2011.2167319

12. Ahmadi MN, Clare PJ, Katzmarzyk PT, et al. Vigorous physical activity, incident heart disease, and cancer: how little is enough? Eur Heart J 2022 doi: 10.1093/eurheartj/ehac572 [published Online First: 20221027]

13. Saint□ Maurice PF, Troiano RP, Matthews CE, et al. Moderate to Vigorous Physical Activity and All□ Cause Mortality: Do Bouts Matter? Journal of the American Heart Association 2018;7(6):e007678. doi: 10.1161/jaha.117.007678

14. Craig CL, Marshall AL, Sjöström M, et al. International physical activity questionnaire: 12-country reliability and validity. Medicine & science in sports & exercise 2003;35(8):1381–95.

15. Khurshid S, Al-Alusi MA, Churchill TW, et al. Accelerometer-derived “weekend warrior” physical activity and incident cardiovascular disease. JAMA 2023;330(3):247–52.

16. Sagelv EH, Hopstock LA, Morseth B, et al. Device-measured physical activity, sedentary time, and risk of all-cause mortality: an individual participant data analysis of four prospective cohort studies. British Journal of Sports Medicine 2023;57(22):1457–63.

17. Hanlon P, Nicholl BI, Jani BD, et al. Frailty and pre-frailty in middle-aged and older adults and its association with multimorbidity and mortality: a prospective analysis of 493 737 UK Biobank participants. The Lancet Public Health 2018;3(7):e323–e32.

18. Tarp J, Hansen BH, Fagerland MW, et al. Accelerometer-measured physical activity and sedentary time in a cohort of US adults followed for up to 13 years: the influence of removing early follow-up on associations with mortality. International Journal of Behavioral Nutrition and Physical Activity 2020;17(1) doi: 10.1186/s12966-020-00945-4

19. Hamer M, Bauman A, Bell JA, et al. Examining associations between physical activity and cardiovascular mortality using negative control outcomes. Int J Epidemiol 2019;48(4):1161–66. doi: 10.1093/ije/dyy272

20. Michel J-P, Leonardi M, Martin M, et al. WHO’s report for the decade of healthy ageing 2021–30 sets the stage for globally comparable data on healthy ageing. The Lancet Healthy Longevity 2021;2(3):e121–e22.

21. Dipietro L, Al-Ansari SS, Biddle SJH, et al. Advancing the global physical activity agenda: recommendations for future research by the 2020 WHO physical activity and sedentary behavior guidelines development group. International Journal of Behavioral Nutrition and Physical Activity 2020;17(1) doi: 10.1186/s12966-020-01042-2

22. Ramírez-Vélez R, Hernández-Quiñones PA, Tordecilla-Sanders A, et al. Effectiveness of HIIT compared to moderate continuous training in improving vascular parameters in inactive adults. Lipids in Health and Disease 2019;18(1):1–10.

23. Allison MK, Baglole JH, Martin BJ, et al. Brief Intense Stair Climbing Improves Cardiorespiratory Fitness. Med Sci Sports Exerc 2017;49(2):298–307. doi: 10.1249/MSS.0000000000001188

24. Lee I-M, Keadle SK, Matthews CE. Fitness Trackers to Guide Advice on Activity Prescription. JAMA 2023

25. Strain T, Wijndaele K, Dempsey PC, et al. Wearable-device-measured physical activity and future health risk. Nature Medicine 2020;26(9):1385–91. doi: 10.1038/s41591-020-1012-

26. Stamatakis E, Owen KB, Shepherd L, et al. Is Cohort Representativeness Passe? Poststratified Associations of Lifestyle Risk Factors with Mortality in the UK Biobank. Epidemiology 2021;32(2):179–88. doi: 10.1097/EDE.0000000000001316

